# Human genetic evidence enriched for side effects of approved drugs

**DOI:** 10.1101/2023.12.12.23299869

**Authors:** Eric Vallabh Minikel, Matthew R. Nelson

## Abstract

Safety failures are an important factor in low drug development success rates. Human genetic evidence can select drug targets causal in disease and enrich for successful programs. Here, we sought to determine whether human genetic evidence can also enrich for labeled side effects (SEs) of approved drugs. We combined the SIDER database of SEs with human genetic evidence from genome-wide association studies, Mendelian disease, and somatic mutations. SEs were 2.0 times more likely to occur for drugs whose target possessed human genetic evidence for a trait similar to the SE. Enrichment was highest when the trait and SE were most similar to each other, and was robust to removing drugs where the approved indication was also similar to the SE. The enrichment of genetic evidence was greatest for SEs that were more drug specific, affected more people, and were more severe. There was significant heterogeneity among disease areas the SEs mapped to, with the highest positive predictive value for cardiovascular SEs. This supports the integration of human genetic evidence early in the drug discovery process to identify potential SE risks to be monitored or mitigated in the course of drug development.

## Introduction

Safety issues are a major contributor to drug candidate failure^1^. The causal evidence of human genetics between drug targets and phenotypic outcomes can provide insights into potential on-target safety liabilities of drug candidates before development has even begun^2^. There are many anecdotes of adverse events supported by genetic evidence^2^ and drug tolerability supported by the lack of negative consequences of gene loss of function in humans^3^. Methodical studies have shown that drug targets with genetic evidence related to a given organ system are more likely to exhibit side effects (SEs) in that organ system^4^, and that drugs with SEs are more likely to bind off-target proteins with related Mendelian diseases^5^. Phenome-wide association studies^6^, curation of loss-of-function variants^7^, and studies investigating the clinical and molecular consequences of rare homozygous loss-of-function participants^8^ been used to evaluate potential on-target liabilities. Yet support for the predictive value of genetic evidence for SEs remains limited.

We recently revisited our prior observation that human genetic evidence supporting the causal relationship between the protein target and indication of a drug, showing that it increases clinical success rates by a factor of 2.6^9^. In so doing, we established a method for mapping human genetically studied traits to drug indications via a similarity matrix based on Medical Subject Headings (MeSH) terms. We used the same approach here to link genetic evidence to SEs and determine whether they are similarly enriched. Assigning quantitative similarity scores allowed us to test the sensitivity of such enrichment to potential confounders. Finally, we estimated the positive predictive value of human genetic evidence, examined the effects of SE frequency, specificity, and severity, and compared the value of genetically-informed predictions across different types of SEs.

## Results

The primary challenge in this research is establishing relationships between drug targets and reported SEs. There is a paucity of systematic data about statistically enriched SEs observed in clinical development. We chose to focus on SEs captured in drug labels and package inserts, limiting us to approved drugs as captured in Side Effect Resource (SIDER)^10,11^, to which we joined our database of human genetic evidence^9^. This resulted in 2,094 unique SEs (MeSH terms), and 567 unique drugs with at least 1 human target, 1 SE and 1 approved indication, or 1,187,298 possible drug-SE pairs (Table S1-S4). Of these possible pairs, the SE was observed (reported in the drug label) for 64,481, yielding an overall base rate (prior probability of an SE being observed for a given drug) of 5.4%.

The primary analysis of interest is the relationship between drug-SE pairs and the presence of genetic evidence between the gene encoding the drug target and a trait similar to the SE. At a trait-SE similarity threshold ≥0.9, we found this strongly enriched (OR = 2.3, 95% CI = 2.2-2.5, Fisher’s exact test). We explored several possible confounders that could affect this observed enrichment (Figure S1A, Table S5). We restricted our analysis to drug-SE pairs where the SE has been studied genetically as well as removing drug-SE pairs where the drug is approved for an indication similar to the SE. This had a modest effect, reducing the OR to 2.0 (95% CI = 1.8-2.1) in combination (Figure S1B, Table S6). We retained both filters for all subsequent analyses. Examining the sensitivity of this enrichment to our SE-trait similarity threshold, we observed ORs above 1 down to 0.2, perhaps recapitulating the previously reported enrichment at the level of organ system^4^ (Figure 1A, Table S7). Varying the threshold for removal of SEs with a similar indication had minimal impact, except at very low thresholds where a large majority of data were removed (Figure 1B, Table S8). These results support the ≥0.9 similarity thresholds used for both metrics. We next examined whether the source of genetic evidence had any influence on this enrichment (Figure 1C, Table S9), and found little difference, though germline oncology evidence had the highest levels of enrichment (Figure S2, Table S10-S11).

**Figure 1.**
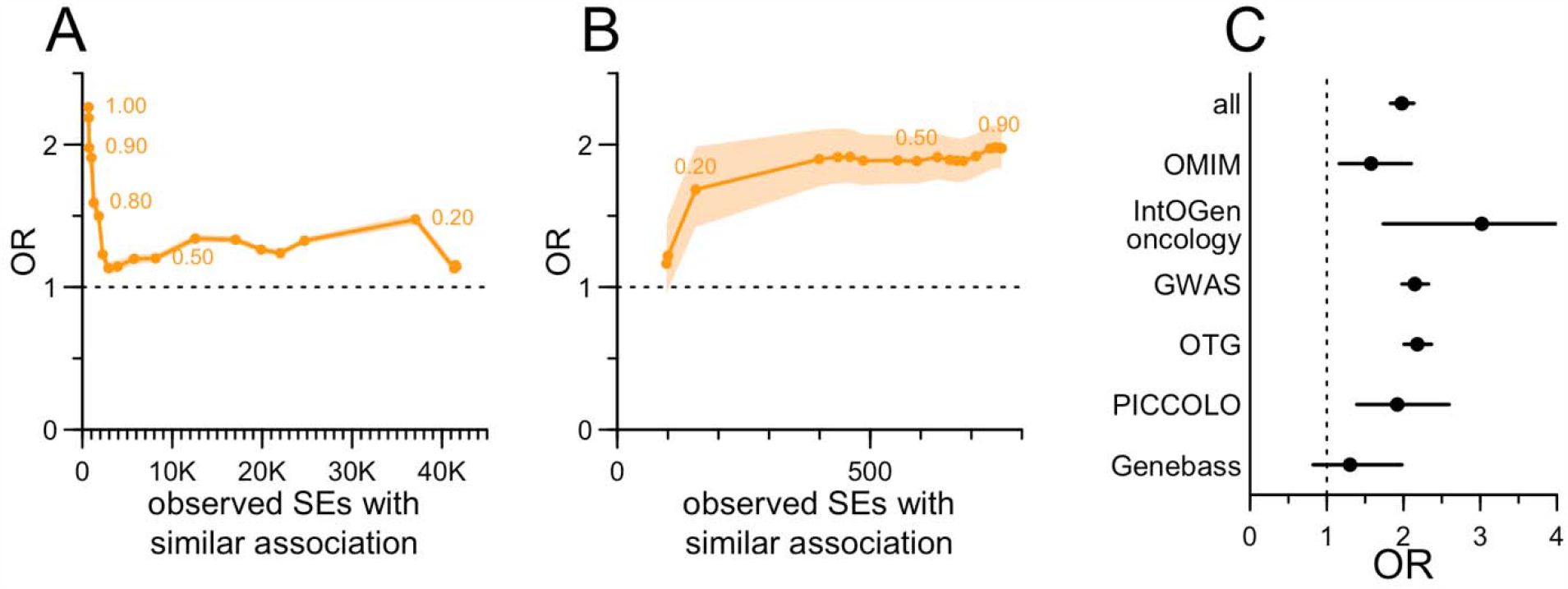
Predictive value of human genetic evidence for labeled drug side effects. **A)** Sensitivity of OR to the threshold for similarity of SE to genetically studied traits; selected similarity thresholds are annotated, resulting number of drug-SE pairs with similar genetic evidence are shown on the x axis and OR is shown on the y axis. Threshold for removal of SEs with similar indications is fixed at 0.9. **B)** As in A, but here the threshold for SE-trait similarity is fixed at 0.9 while the similarity threshold between the SE and approved indications is varied. **C)** OR for several sources of genetic evidence contributing to this study.

SIDER provides several SE modifiers extracted from the text, including frequency as numerical estimates or descriptive terms and whether the SEs were supported by placebo or non-placebo-controlled studies. Where numerical frequencies were available, we found that genetic evidence enrichment increases with increasing SE frequency (P = 7.4e-8, binomial logit, Figure 2, Table S12-S13). The effect was more variable when analyzed using descriptive frequency terms ranked by their reported perceived numeric values^12^ (P = 0.057 for the linear term in a binomial logit, Figure 2, Table S14-S16), though the highest point estimates were for SEs described as common or very common. Estimates of enrichment were higher for SEs backed by placebo-based evidence, but that difference was not statistically significant (P = 0.079, binomial logit, Figure 2, Table S17-S18).

**Figure 2.**
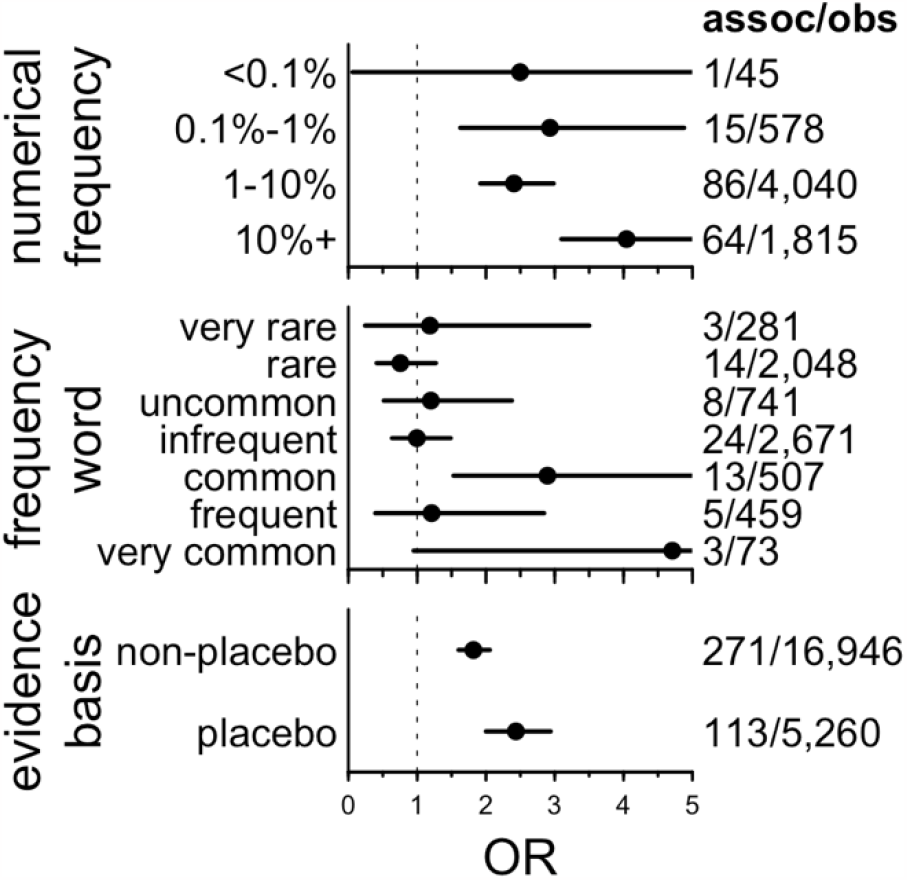
Impact of SE modifiers on genetic evidence enrichment. Because frequency and evidence basis are only defined for observed side effects, the OR indicated here is the enrichment of genetic evidence conditioned on an SE being observed with the indicated modifier. The assoc/obs fraction indicates in the denominator the number of drug-SE combinations observed with the indicated properties and in the numerator the number of those that have genetic evidence. The ordering of the frequency words is based on ref. ^12^.

We further explored enrichment of genetic evidence based on the number of different drugs for which an SE was reported, without respect to the underlying target gene. We found it was strongest for SEs observed for 2-9 drugs, and decreased as the number of drugs increased (Figure 3A, Table S19). This poses a challenge for practical utility of human genetics in predicting SEs. The SEs that are most informed by genetic evidence are more drug-specific, and highly drug-specific SEs necessarily have a low base rate, resulting in relatively low predictive values (PPVs; probability of observing an SE given genetic evidence, Figure 3B, Table S19).

**Figure 3.**
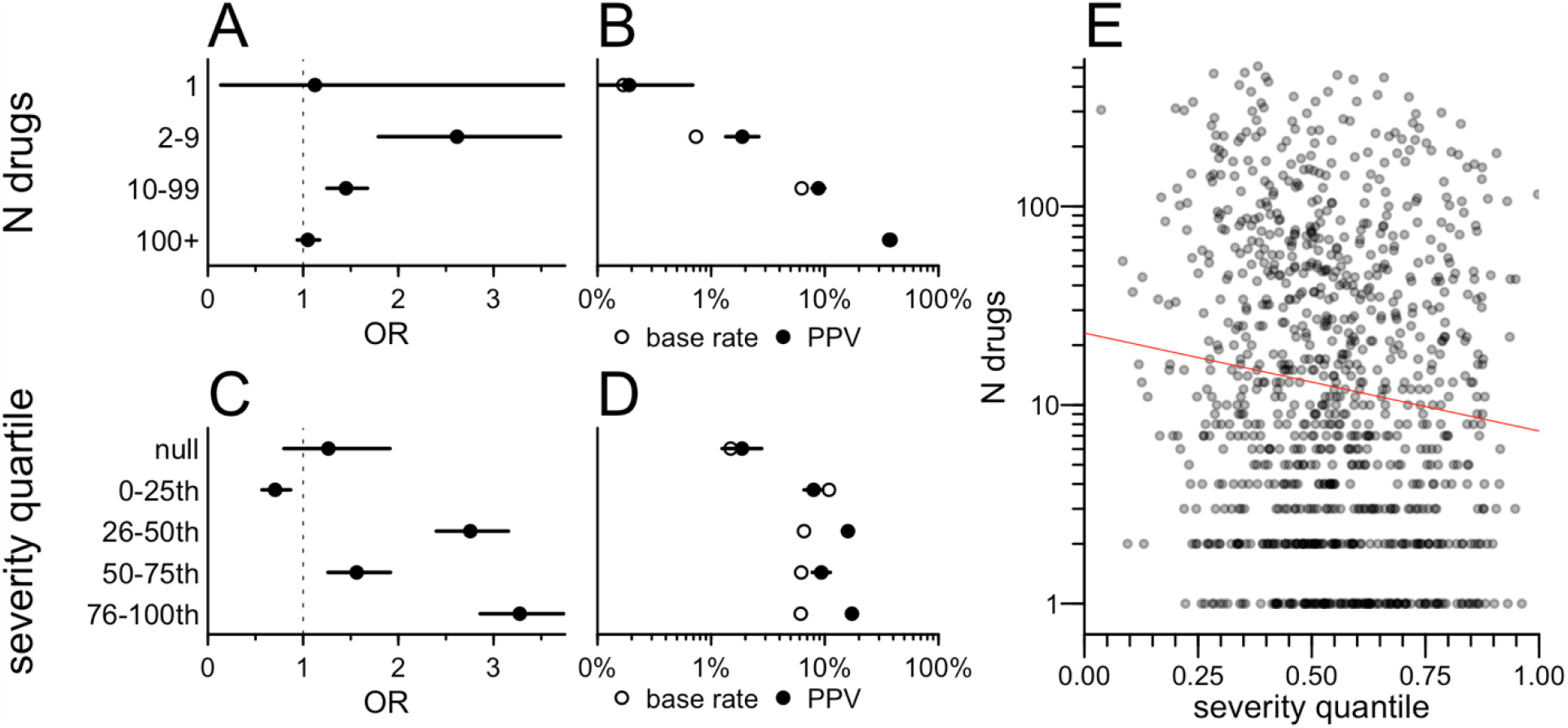
Relationship between SE specificity and severity, and the predictive value of genetic evidence. **A)** OR for enrichment of genetic evidence, binned by the number of drugs for which SE was observed. The unit of analysis is drug-SE pairs; thus, the number of observed drug-SE pairs is necessarily higher for those SEs observed for a larger number of drugs, hence the tighter confidence intervals in the “100+” bin compared to the “1” bin. **B)** Base rate (proportion of drugs reporting the SE) and positive predictive value (proportion of drugs with genetic evidence for the SE) binned as in A. Note that the higher base rate for those SEs observed for a larger number of drugs is tautological. **C)** OR by quartiles of SE severity. **D)** Base rate and positive predictive value binned as in C. **E)** Correlation between severity and specificity; each point is an SE, x axis indicates its severity quantile and y axis indicates the number of drugs for which it is observed.

Using crowdsourced severity rankings of the observed SEs^13^, we found that OR was also positively, though non-monotonically, associated with SE severity (P = 3.0e-23, binomial logit; Figure 3C, Table S20-S21), while the base rate was slightly lower for the more severe SEs (Figure 3D, Table S20). SE severity and specificity were themselves correlated (P = 2.1e-4, log-linear regression; Figure 3E, Table S22) with more severe SEs tending to be observed for fewer drugs.

To better understand the value of genetic evidence on SE risk, we next binned SEs by top level disease headings of the MeSH ontology (Figure 4, Table S23), revealing substantial heterogeneity in OR (P < 1e-15, CMH test; Figure 4A). Endocrine-related SEs, for instance, had the largest effect (OR = 6.5) with a low base rate (1.9%) and a PPV of 10.5% (Figure 4B), and were moderately severe (Figure 4C, Table S23). In contrast, cardiovascular SEs had a combination of high base rate and high OR resulting in the highest PPV (27.7%), and tended to be relatively severe. PPV and OR were not significantly correlated across SE areas (ρ = 0.38, P = 0.14, Spearman) nor between severity and base rate (ρ = -0.24, P = 0.36, Spearman). These findings were broadly consistent when we removed drugs where an approved indication fell within the same area as the SE (Figure S2, Table S23).

**Figure 4.**
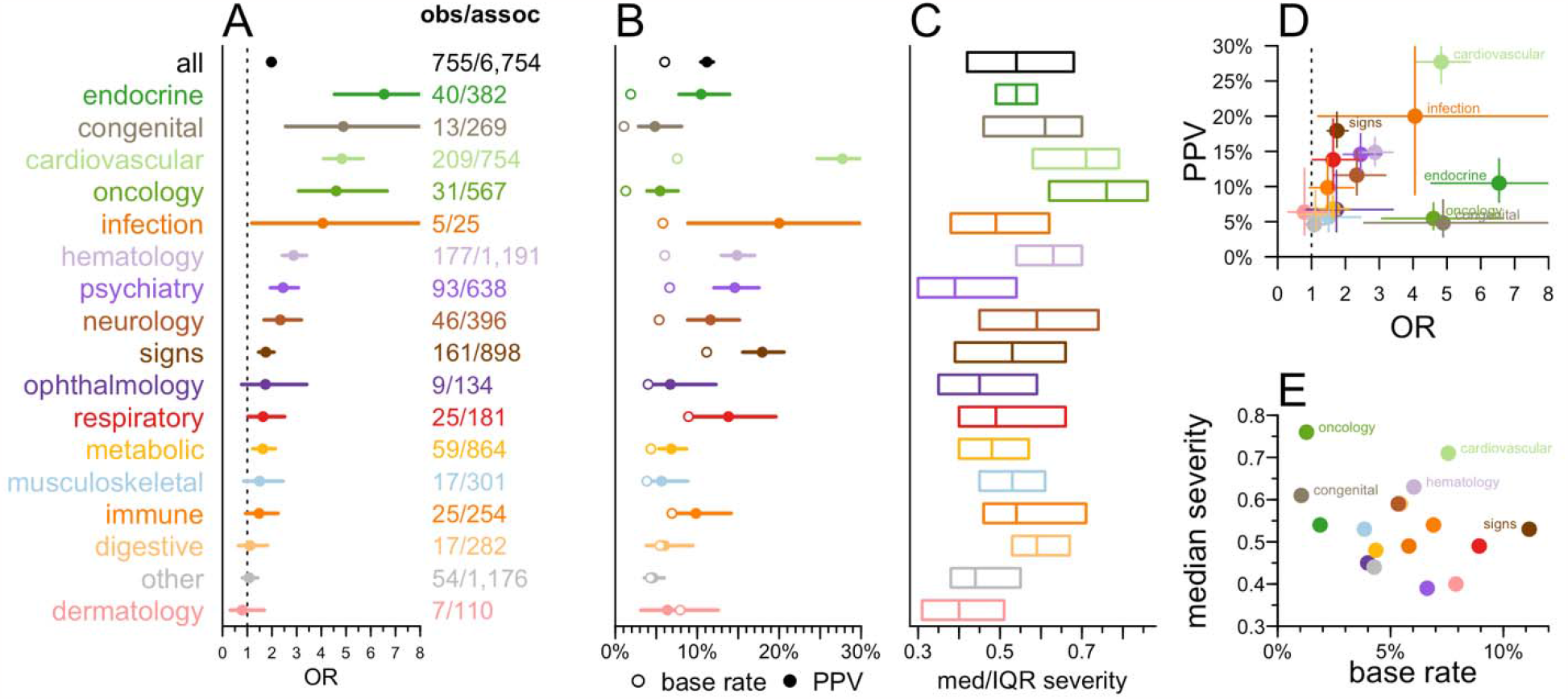
Utility of human genetic evidence for predicting side effects by affected function or organ system. **A)** OR binned by the SE’s top-level heading within the Medical Subject Headings (MeSH) ontology. Fractions indicate the number of drug-SE pairs with genetic evidence (denominator) and of those, the number that were observed (numerator). **B)** Base rate (mean proportion of drugs with the side effect) and positive predictive value (proportion of drugs with genetic evidence that exhibit the SE) were binned as in A. **C)** Median and interquartile range (IQR) of severity quantiles for SEs in each bin. **D)** Positive predictive value (PPV) vs. OR across and **E)** median severity vs. base rate across SE areas.

## Discussion

An important limitation of our analysis is restriction to SEs for approved drugs. The requirement for drugs to exhibit a favorable risk/benefit balance to achieve approval presumably constrains SEs to be less frequent or less severe than would be the case for drugs in clinical development. It is possible that human genetic evidence has different predictive value for SEs observed in trials that could result in termination than for labeled for approved drugs.

Our results demonstrate that human genetic evidence identifies on-target drug mechanisms that are at increased risk for SEs among approved drugs. Genetic evidence of cardiovascular effects were particularly predictive of SEs, with a PPV of nearly 30% and relatively high reported severity. These results support the use of genetic evidence to identify potential SE risks that can be monitored and potentially mitigated during drug discovery and clinical development.

## Online Methods

### Side effects

Side effect data were obtained from the SIDER database^11^ (v4.1). Citeline Pharmaprojects^14^ and DrugBank^15^ were parsed as described^7,9,16^, and SIDER drug names were mapped to Pharmaprojects indications using text matches to Pharmaprojects drug name synonyms, or, by mapping first to DrugBank using either ATC codes or name matches to obtain CAS numbers, and then looking up CAS numbers in Pharmaprojects; the proportion of drugs mapped by various approaches is provided in Table S2. Pharmaprojects matches were used to obtain human gene targets and MeSH terms for approved indications as described^9^. Side effects were mapped to MeSH terms using UMLS MedDRA – MeSH mapping, exact term and substring match to UMLS and MeSH, and manual curation; the proportion of terms mapped by various approaches is provided in Table S3. We removed drugs that were duplicates, lacked an annotated human target, an annotated approved indication, or were unmappable (Table S4). Severity rankings were taken from a crowdsourcing study^13^. Ordering of frequency terms was based on numerical values determined empirically with human participants^12^.

### Human genetic evidence

We used human genetic evidence from OMIM^17^, Open Targets Genetics^18^, PICCOLO^19^, Genebass^20^, and IntoGen^21^; the filtering, aggregation, and MeSH mapping of this dataset has been described^9^. Open Targets Genetics gene mappings were filtered to those with ≥50% of the total share of locus-to-gene (L2G) score assigned to any gene. As before^9^, we considered a trait to have been studied genetically if there was at least 1 OMIM or IntOGen gene, or at least 3 unlinked GWAS associations for a trait with ≥0.8 similarity.

### Similarity mapping

Similarity between MeSH terms for SEs, genetically associated traits, and drug indications was computed using combined Lin and Resnik similarity scores^22,23^ as described^9^. MeSH terms were further mapped onto MeSH top level headings as described^9^.

### Target enrichment

To test whether a particular side effect was enriched among drugs with a particular target, we performed a Fisher exact test on the contingency table of drugs with and without the target of interest, with and without the side effect of interest reported. Target-SE combinations yielding an odds ratio ≥2 and a nominal P value <0.01 were considered to be enriched. We note that the power to detect such enrichment is confounded with the number of drugs sharing a particular target and with the number of drugs for which the side effect is reported. Moreover, drugs sharing a target may also be of the same chemical class and may therefore share off-target liabilities. In consideration of these limitations, we considered target enrichment among our candidate variables in Figure 1A but did not use this metric in ensuing analyses.

### Models and statistics

Analyses utilized custom scripts in R 4.2.0. The primary metric in this analysis — whether an SE is more likely to be observed when there was genetic evidence — was computed as an odds ratio (OR) from a Fisher exact test on the 2 × 2 contingency table of drugs with and without an SE, whose targets do or do not have a genetic evidence. Following the findings of Figure 1A-B, this was computed after removing drugs with an indication similar to the SE, and after removing SEs not studied genetically. The shaded areas for curves and error bars in forest plots represent the 95% confidence intervals from this Fisher test. Binomial logit models used the SE’s occurrence as the dependent variable, and genetic evidence and the variable of interest (for instance, SE severity) as independent variables, with interaction terms — in R, glm(observed ∼ sim_assoc * severity, family=‘binomial’). The Cochran-Mantel-Haenszel (CMH) test for heterogeneity was performed across these 2 × 2 contingency tables for each MeSH area. For analysis of attributes that are only defined when the SE is observed (frequency and placebo status in Figure 2), the Fisher test was based on the 2 × 2 contingency table of drugs that do or do not have an SE *with the stated attributes* (for instance, frequency >10% in patients), whose targets do or do not have genetic evidence. In such instances, binomial logit models used presence of genetic evidence as the dependent variable, and the variable of interest (for instance, numerical frequency) as the independent variable: e.g. glm(sim_assoc ∼ frequency, family=‘binomial’). Frequency terms were treated alternatively as ordinal variables, resulting in terms for linear and higher-order terms, or as numerical variables with the term’s rank as the numerical value, resulting in only a linear term. Base rate, or an SE’s drug specificity, was defined as the proportion of drug-SE pairs for which the SE was observed.

Positive predictive value (PPV) was defined as the number of drug-SE pairs where the SE was observed and was supported by genetic evidence, divided by the total number of drug-SE pairs where the target had genetic evidence. Linear regression for specificity versus severity used the logarithm of the number of drugs for which an SE was observed, in R: lm(log(n_drugs) ∼ severity). Correlations across MeSH areas were tested using Spearman rank correlations. All tests were two-sided, and P values less than 0.05 were considered to be nominally significant.

## Data Availability

All data produced in the present work are contained in the manuscript, the supplementary tables, or the public GitHub site at https://github.com/ericminikel/genetics_side_effects/

## SOURCE CODE AVAILABILITY AND DATA AVAILABILITY

An analytical dataset and source code will be made available at https://github.com/ericminikel/genetics_side_effects/ and will be sufficient to reproduce all figures and statistics herein.

## SUPPLEMENT

**Figure S1.**
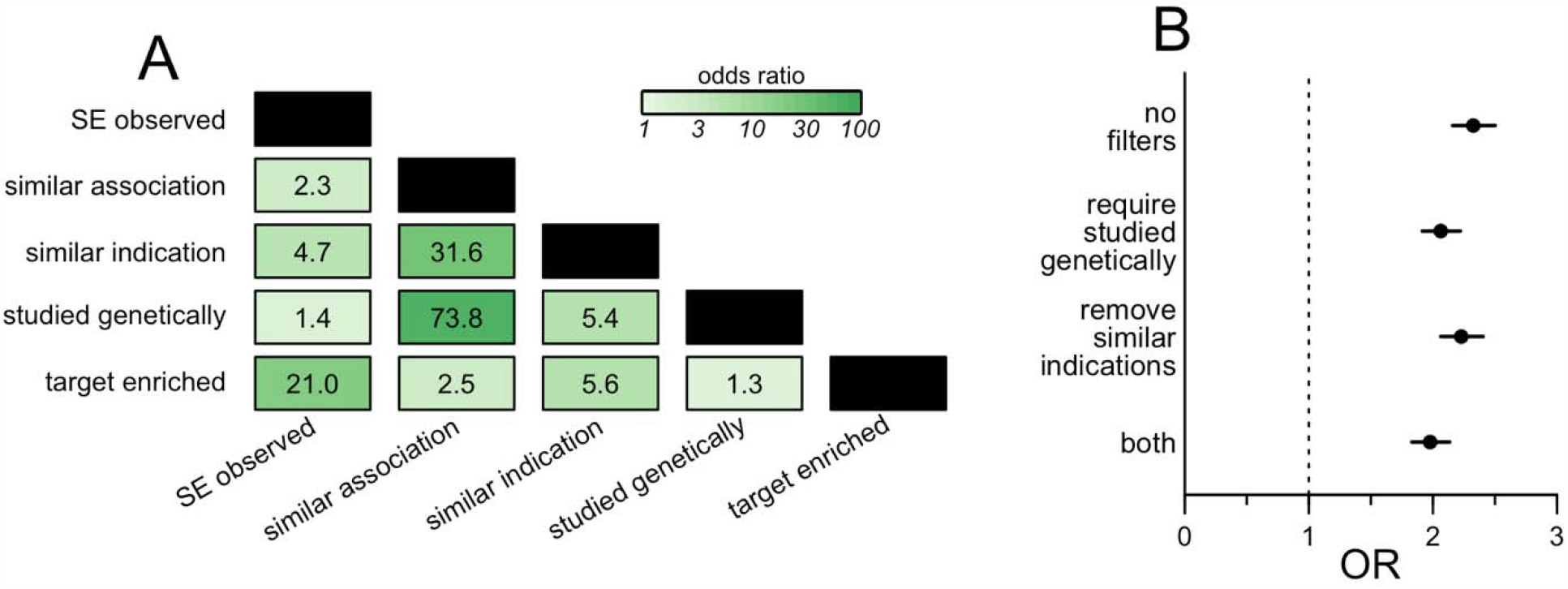
Examination of possible confounders and establishment of metric used throughout. **A)** Correlogram showing the odds ratios (ORs) by Fisher exact test for enrichment of all combinations of properties (Table S1, Table S5, Methods) evaluated in the dataset. **B)** OR for enrichment of genetic evidence vs. SE observed, with the indicated filters applied.

**Figure S2.**
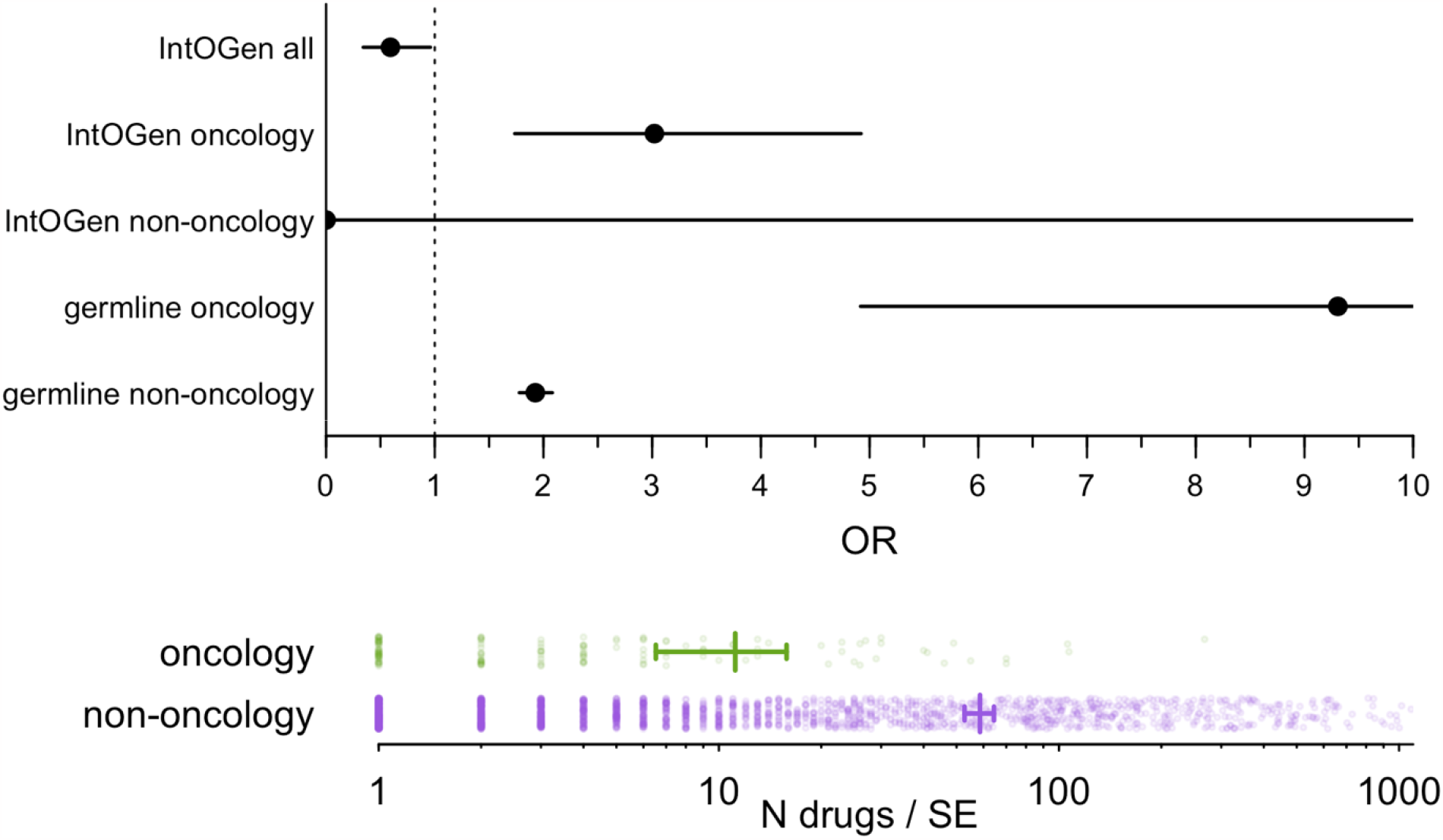
Breakdown of evidence sources for oncology. **A)** Forest plot of OR by source of evidence (IntOGen somatic evidence vs. all sources of germline evidence) versus oncological and non-oncological SEs. **B)** Drug specificity of oncological and non-oncological SEs. IntOGen overall has an OR < 1 because its somatic evidence are almost exclusively similar to oncological SEs, which are more drug-specific than non-oncological SEs. Thus, the IntoGen OR for oncology only is shown in Figure 1. Germline evidence appears to have a higher OR than somatic evidence for oncology.

**Figure S3.**
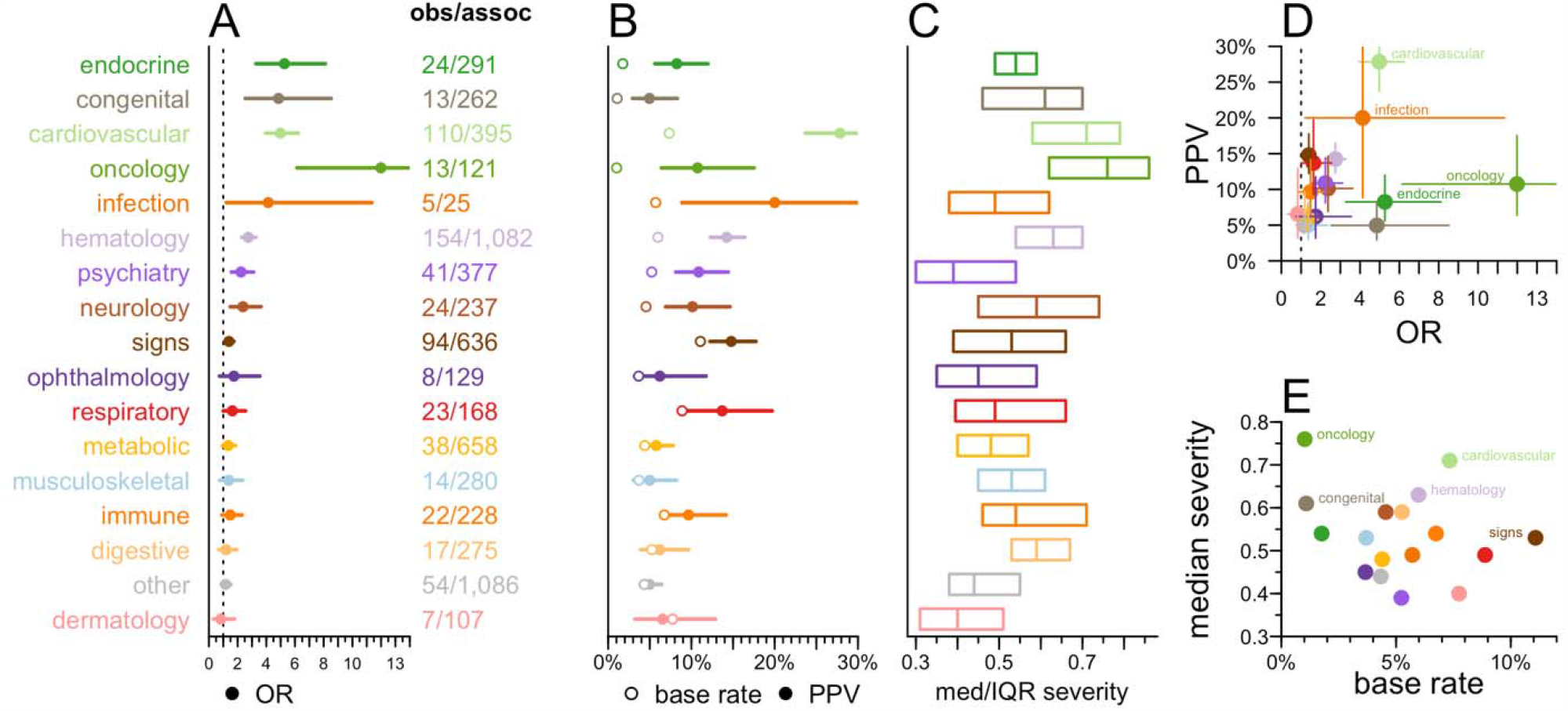
Breakdown by side effect area. As Figure 4, but within each MeSH area, any drug with any indication in that area is removed.

Tables S1-S23 are provided as a separate Excel file and are available online.

